# Olfactory impairment is related to tau pathology and neuroinflammation in Alzheimer’s disease

**DOI:** 10.1101/2020.08.31.20183558

**Authors:** Julia Klein, Xinyu Yan, Aubrey Johnson, Zeljko Tomljanovic, James Zou, Krista Polly, Lawrence S. Honig, Adam M. Brickman, Yaakov Stern, D.P. Devanand, Seonjoo Lee, William C. Kreisl

## Abstract

**Background:** Olfactory impairment is evident in Alzheimer’s disease (AD), however, its precise relationships with clinical biomarker measures of tau pathology and neuroinflammation are not well understood.

**Objective:** To determine if odor identification performance measured with the University of Pennsylvania Smell Identification Test (UPSIT) is related to *in vivo* measures of tau pathology and neuroinflammation.

**Methods:** Participants were selected from an established research cohort of adults aged 50 and older who underwent neuropsychological testing, brain MRI, and amyloid PET. Fifty-four participants were administered the UPSIT. Forty-one underwent ^18^F-MK-6240 PET and fifty-three underwent ^11^C-PBR28 PET to measure tau pathology and neuroinflammation, respectively. Twenty-three participants had lumbar puncture to measure CSF concentrations of total tau (t-tau), phosphorylated tau (p-tau) and β-amyloid (Aβ_42_).

**Results:** Low UPSIT performance was associated with greater^18^F-MK-6240 binding in medial temporal cortex, hippocampus, middle/inferior temporal gyri, inferior parietal cortex and posterior cingulate cortex (p < 0.05). Similar relationships were seen for ^11^C-PBR28. These relationships were primarily driven by amyloid-positive participants. Lower UPSIT performance was associated with greater CSF concentrations of t-tau and p-tau (p < 0.05). Amyloid status and cognitive status exhibited independent effects on UPSIT performance (p < 0.01).

**Conclusions:** Olfactory identification deficits are related to extent of tau pathology and neuroinflammation, particularly in those with amyloid pathophysiology. The independent association of amyloid-positivity and cognitive impairment with odor identification suggests that low UPSIT performance may be a marker for AD pathophysiology in cognitive normal individuals, although impaired odor identification is associated with both AD and non-AD related neurodegeneration.

**NCT Registration Numbers:** NCT03373604; NCT02831283

## INTRODUCTION

Olfactory impairment is observed early in Alzheimer’s disease (AD) [1-4] and is thought to occur due to anatomical overlap of the regions involved in olfaction and early AD pathology. Olfactory bulb neurons project directly to limbic regions of the brain for olfactory processing [5, 6]. These regions, including the transentorhinal cortex and medial temporal lobes, are known to be involved in early tau pathological changes of AD and correspond with Braak stages 1-2 [5-7]. Tau pathology in the olfactory bulb continues to increase with severity of AD, which provides a possible explanation for the progression of odor impairment that occurs with AD advancement [8].

Olfactory impairment observed in AD can be quantified with the University of Pennsylvania Smell Identification Test (UPSIT). UPSIT scores appear to correlate with measures of entorhinal cortex volume on MRI [9, 10]. Large community cohort studies have demonstrated that low UPSIT scores predict cognitive decline in cognitively normal elders and patients with mild cognitive impairment (MCI) [3, 11, 12]. These studies have also shown that UPSPIT performance is inversely related to performance on neuropsychological testing [12].

Several studies have investigated the relationships between UPSIT and *in vivo* measures of AD pathology, particularly amyloid. Some studies have demonstrated modest relationships between UPSIT performance and amyloid deposition on PET [9, 10]. Only one published study has examined the relationship between odor identification and PET measures of tau pathology, with results indicating that binding with the tau radioligand ^18^F-AV-1451 negatively correlated with UPSIT performance in cognitively normal adults, adults with subjective cognitive decline, and MCI patients [13]. However, that study did not include AD patients. One study evaluated the relationship between odor identification and CSF measures of tau pathology, demonstrating that low UPSIT performance was associated with elevated CSF tau [14].

Neuroinflammation is also associated with AD pathology and cognitive decline [15] and can be quantified using PET radioligands, such as ^11^C-PBR28, that bind the 18 kDa translocator protein (TSPO), a marker of immune activation. To our knowledge, no study has evaluated the relationship between odor identification and neuroinflammation.

We sought to determine the relationship between odor identification and neuroinflammation, measured by ^11^C-PBR28 PET. We further evaluated relationships between odor identification and tau pathology using PET imaging with ^18^F-MK-6240, a highly specific radioligand for phosphorylated tau, and CSF concentrations of total tau (t-tau) and phosphorylated tau (p-tau), and the relationship between odor identification and amyloid pathology using CSF concentrations of β-amyloid (Aβ_42_). We hypothesized that worse performance on odor identification testing would be associated with higher PET measures of neuroinflammation, and higher PET and CSF-biomarker measures of tau pathology, particularly in regions of early AD pathology, namely medial temporal lobe structures.

## METHODS

### Participant selection

Adults aged 50 years and older were recruited from Columbia University Irving Medical Center (CUIMC) Aging and Dementia clinic, the Columbia University Alzheimer’s Disease Research Center, other research cohorts at CUIMC or self-referral to establish the initial research cohort for a larger study (K23AG052633, PI = Kreisl). A subset of seventy-eight adults from the initial research cohort was considered for inclusion into this study.

All seventy-eight participants underwent an initial screening that included routine history and physical, neurological examination, routine laboratory tests, *TSPO* genotyping, neuropsychological evaluation and brain MRI. Screening measures were performed to exclude any participants with significant medical or psychiatric illness, cortical infarcts on brain MRI, and use of immunosuppressant medication. After screening, seventeen subjects were excluded from continuing participation in the study, including eight due to low affinity *TSPO*, three due to lab exclusions and six withdrawals (Supplementary Figure 1).

Neuropsychological evaluation including the Mini Mental State Examination, [16] Selective Reminding Test-Delayed Recall (SRT-DR), [17] Trail Making Test Parts A and B, and Category and Phonemic Fluency. These tests were selected to capture performance of specific cognitive domains, while the MMSE provided a global representation of cognition. The SRT-DR tested shor-term memory, Trail Making Test Part A tested psychomotor functioning, Trail Making Test Part B tested executive functioning and Category and Phonemic Fluency tested language fluency. All cognitive test scores were transformed into z-scores using age-, sex- and education-adjusted normative data provided by the National Alzheimer’s Coordinating Center. All participants were assigned a Clinical Dementia Rating scale score (CDR) by a clinician based on history, examination, and neuropsychological test results. Only participants with a CDR score ≤1 (i.e. normal, mild cognitive impairment, or mild AD) were eligible, so this study could focus on pathological changes in early stages of AD, and to ensure that participants were able to complete study procedures.

Participants were defined as either cognitively normal or cognitively impaired based on history and cognitive examination. To qualify as cognitively impaired, participants had to have a primary memory complaint and meet clinical criteria for amnestic mild cognitive impairment (MCI) [18] or AD [19]. Participants who met clinical criteria for a non-AD neurodegenerative condition (e.g., dementia with Lewy bodies, vascular dementia, Parkinson’s disease, corticobasal degeneration, progressive supranuclear palsy, or frontotemporal dementia) were excluded. To qualify as cognitively normal, participants had to have no cognitive complaints and have absence of clinically significant cognitive impairment based on history and neuropsychological evaluation.

### TSPO affinity determination

Blood samples were collected from all participants at the initial screening visit to utilize genomic DNA to genotype the rs6971 polymorphism using a TaqMan assay [20]. Participants from the original cohort (n = 78) that were homozygous for the low affinity allele were excluded from continuing to participate in the study (n = 8) (Supplementary Figure 1), as those with this *TSPO* genotype show negligible binding on ^11^C-PBR28 PET [21].

### Amyloid PET Imaging

The sixty-one participants who met inclusion criteria after initial screening procedures had PET imaging with ^18^F-florbetaben (FBB) to determine amyloid status in a Siemens Biograph64 mCT/PET scanner at the CUIMC Kreitchman PET center (target dose: 8.1 mCi; 4×5 min frames), with a low-dose CT scan for attenuation correction. FBB images were acquired 50-70 minutes post-injection. All PET data were corrected for radioactive decay, attenuation of annihilation photons, scanner deadtime and normalization, and random and scatter events. Reconstructed FBB images were averaged to create a single static image for each participant. Amyloid status was determined by a binary visual read by an experienced neurologist (WCK), blinded to the participant diagnosis, according to established methods [22]. To validate the visual reads, we determined a SUVR cutoff of 1.27 for FBB as defined by the minimum among the visually amyloid-positive participants (Supplementary Figure 2). Using this cutoff, we found concordance in amyloid status determination between visual reads and use of SUVR in 58 of 61 participants (95.1%). The three discordant cases were then reviewed by a second trained and experienced reader (AJ), blind to diagnosis and the first reader’s interpretations, who agreed with the first reader on all thee visual interpretations. Therefore, we used the visual read results as the determinant for amyloid positivity or negativity. Studies have indicated that visual assessments perform similarly to SUVR cutoffs in interpreting amyloid status with FBB scans [23].

### Odor Identification Test Administration and Scoring

The 40-item UPSIT was administered by a trained technician on the same day as either the ^18^F-MK-6240 or ^11^C-PBR28 scan. For each of the 40 items on the UPSIT, the participants were provided with an odorant embedded in a microcapsule that could be scratched and smelled. They were instructed to choose from four distinct answer choices. The test was scored between 0 (no odors correctly identified) and 40 (all odors correctly identified). Because there is a 25% chance of guessing each odorant correctly, scores of 10 or below are consistent with anosmia and therefore were excluded from the analysis.

UPSIT was completed in 55 participants who had FBB PET. Four participants reported history of anosmia and did not have UPSIT performed. Two were unable to complete UPSIT. One participant who scored a 10 on the UPSIT was therefore excluded from analysis, leaving 54 participants with useable UPSIT data. Other known factors contributing to hyposmia such as smoking and current upper respiratory infection were considered, however, no participants were smokers or experiencing upper respiratory symptoms at the time of testing.

### MRI Acquisition and Processing

T1-weighted MRI scans (160 slice 1 mm resolution, 256 x 200 voxel count) were acquired for all participants on a 3T Phillips Achieva MRI machine at CUIMC. Using PMOD 3.8 (PMOD Technologies), the T1 MR images were segmented and normalized to standard space. The Hammers-N30R83-1MM atlas was used to define regions of interest (ROIs), which were then consolidated into 10 volume-weighted ROIs. These ROIs included prefrontal cortex (middle frontal gyrus, superior/inferior frontal gyrus, posterior orbital gyrus); middle and inferior temporal gyri (medial part of anterior temporal lobe, lateral parts of anterior temporal lobe and middle and inferior temporal gyri); superior temporal gyrus (anterior part of superior temporal gyrus, posterior part of superior temporal gyrus); medial temporal cortex (amygdala, parahippocampal gyrus); posterior cingulate cortex; superior parietal lobule; inferior parietal lobule; lingual gyrus; striatum (caudate nucleus and putamen); and cerebellum. ROI volumes were reverse-warped to the participant’s native MRI space and manually corrected, if required. Left and right hippocampi were manually drawn on the native MRI by blinded investigators and the weighted-average volume was used as an ROI distinct from the remainder of the medial temporal cortex (i.e., the PMOD-derived amygdala and parahippocampal gyrus). The volume of each ROI was divided by total intracranial volume to adjust for differences in brain size.

### Tau and Neuroinflammation PET Imaging

Forty-one participants underwent ^18^F-MK-6240 PET imaging to measure tau pathology (target dose: 5 mCi; 6×5 min frames). ^18^F-MK-6240 images were acquired 80-100 min post injection. Fifty-three participants underwent ^11^C-PBR28 PET imaging to measure TSPO (target dose: 20 mCi; 6×5 min frames). ^11^C-PBR28 PET images were acquired 60-90 min post-injection. ^18^F-MK-6240 and ^11^C-PBR28 PET imaging were performed on the same scanner as the FBB scans. Because it is a relatively novel radioligand, PET imaging with ^18^F-MK-6240 was not available at the initiation of this study. Therefore, not all fifty-four participants who completed the UPSIT were able to undergo ^18^F-MK-6240 PET imaging.

### PET Image Processing

^18^F-MK-6240 and ^11^C-PBR28 PET images underwent the same processing steps. Reconstructed images were realigned and then corrected for participant movement with SPM12 (Wellcome Centre for Human Neuroimaging). The PNEURO tool in PMOD 3.8 was then used to coregister PET images into native MRI space and to perform correction for partial volume effects with the region-based voxelwise method [24]. The dynamic frames were then averaged to a single static image and the native MRI space ROIs defined above were applied to the averaged PET image. The concentration of radioactivity of each ROI was divided by the concentration of radioactivity of a reference region to generate standardized uptake value ratios (SUVRs). For ^18^F-MK-6240, inferior cerebellar gray matter was used as a reference region to avoid spill-over into the anterior lobe of the cerebellum from ventral temporal and occipital cortex [25]. For ^11^C-PBR28, the entire cerebellar gray matter was used as a “pseudo-reference” region, as previously validated [23, 26]. For ^18^F-MK-6240 and ^11^C-PBR28, both partial volume-corrected and uncorrected SUVRs were calculated.

### CSF analysis

Lumbar puncture was optional for study participants. Twenty-three participants who had UPSIT also agreed to lumbar puncture and had CSF collected to measure concentrations of t-tau, p-tau (phosphorylated at threonine 181) and Aβ_42_.

Up to 15 cc of CSF was removed using a Sprotte 24G spinal needle, and placed in two 12 cc polypropylene tubes. All samples were centrifuged briefly, aliquoted using polypropylene pipettes within 30 minutes, and stored at −80°C. T-tau, p-tau (181), and Aβ_42_ concentrations were measured using the micro-bead-based multiplex immunoassay, the INNO-BIA AlzBio3 kit (Fujirebio, Ghent, Belgium), on the Luminex platform [27].

### Statistical Analysis

Study participants were grouped based on amyloid status and CDR score into four groups: amyloid-negative controls (CDR = 0), amyloid-positive controls (CDR = 0), amyloid-positive patients (CDR = 0.5-1) and amyloid-negative patients (CDR = 0.5-1). For key characteristics, mean and standard deviations for continuous variables and frequencies for categorical variables were presented by each group. For continuous variables, the difference between groups was compared using analysis of variance (ANOVA) followed by post-hoc pairwise group difference tests, uncorrected for multiple comparisons. To assess the effect of amyloid status and cognitive status on UPSIT performance, we performed a 2-way ANOVA with factors amyloid status and cognitive status, controlling for age, sex, and *TSPO* genotype. Partial eta squared (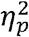) were calculated as effect size measures. Categorical demographic variables (e.g., sex, *TSPO* genotype) were tested for group differences with Chi-squared tests.

Partial correlation analyses evaluated the association between UPSIT total score and ^11^C-PBR28 binding, ^18^F-MK-6240 binding, CSF biomarkers, MMSE scores, and SRT-DR scores, covarying for age and sex (and *TSPO* genotype when applicable). The same partial correlation analyses were performed by amyloid status group (positive *and* negative) separately. For ^11^C-PBR28 binding and ^18^F-MK-6240 binding, partial correlation coefficients (*r_p_*) were computed in each ROI. The p-values of whole group association regarding ^11^C-PBR28 binding, ^18^F-MK-6240 binding, and CSF biomarkers were corrected for multiple comparisons controlling for false discovery rate [28]. Uncorrected p-values are also reported.

All statistical analyses were performed in R, version 3.6.0. Graphs were generated using GraphPad Prism 8. For visualization, residuals were calculated by regressing each variable on age, sex, and *TSPO* genotype.

### Standard protocol approvals, registrations, and patient consents

This study was approved by the Columbia University Irving Medical Center Institutional Review Board. All participants (or their representative) provided informed consent according to the Declaration of Helsinki for participation in the study and for their health information to be used for research purposes.

### Data availability

Anonymized data will be made available upon reasonable request to qualified investigators.

## RESULTS

### Participant Demographics

Fifty-four participants completed screening procedures and ^18^F-FBB PET scan and had UPSIT performed (23 amyloid-positive patients, 9 amyloid-negative patients, 6 amyloid-positive controls, 16 amyloid-negative controls) (Table 1). Forty-one of these participants (16 amyloid-positive patients, 8 amyloid-negative patients, 6 amyloid-positive controls, 11 amyloid-negative controls) underwent ^18^F-MK-6240 PET scan. Fifty-three of these participants (22 amyloid-positive patients, 9 amyloid-negative patients, 6 amyloid-positive controls, 16 amyloid-negative controls) underwent ^11^C-PBR28 PET scan. Twenty-three of these participants also underwent lumbar puncture (12 amyloid-positive patients, 3 amyloid-negative patients, 4 amyloid-positive controls, 4 amyloid-negative controls).

**Table 1.** Descriptive data for participant demographics based on amyloid and cognitive status^a^

|  | **Aβ (+)**<br>**patients**<br>**(n = 23)** | **Aβ (+)**<br>**controls**<br>**(n = 6)** | **Aβ (-)**<br>**patients**<br>**(n = 9)** | **Aβ (-)**<br>**controls**<br>**(n = 16)** | **F-statistic**<br>**(continuous) /**<br>**Chi-Squared**<br>**(categorical)** | **P-value** |
| --- | --- | --- | --- | --- | --- | --- |
| **Age (years)<sup>b</sup>** | 64.7 ± 8.6 | 71.3 ± 4.6 | 74.0 ± 8.0 | 67.8 ± 3.8 | 4.27 | 0.01 |
| **Male/Female** | 19/4 | 3/3 | 6/3 | 5/11 | 10.90 | 0.01 |
| **Education (years)** | 16.9 ± 2.4 | 15.0 ± 2.8 | 16.8 ± 3.5 | 15.8 ± 2.8 | 1.11 | 0.35 |
| **MMSE score<sup>b, c, d</sup>** | 23.6 ± 4.2 | 28.7 ± 2.1 | 27.6 ± 2.3 | 29.4 ± 0.8 | 13.15 | 0.00 |
| **SRT-DR (z-score)<sup>c, d, e, f</sup>** | -3.26 ± 0.65 | +0.26 ± 1.30 | -2.55 ± 0.69 | 0.74 ± 1.05 | 76.92 | 0.00 |
| **TSPO genotype**<br>**(HAB/MAB)** | 12/11 | 2/4 | 5/4 | 11/5 | 2.43 | 0.49 |
| **% Hippocampal**<br>**Volume<sup>c, f</sup>** | 0.87 ± 0.16 | 1.00 ± 0.16 | 0.84 ± 0.18 | 1.05 ± 0.14 | 5.61 | 0.00 |

<sup>a</sup> Thirteen participants did not undergo <sup>18</sup>F-MK-6240 PET and 1 participant did not undergo <sup>11</sup>C-PBR28 PET
<sup>b</sup> Significant difference between Aβ (+) patients and Aβ (-) patients ( $p < 0.05$ )
<sup>c</sup> Significant difference between Aβ (+) patients and Aβ (-) controls ( $p < 0.05$ )
<sup>d</sup> Significant difference between Aβ (+) patients and Aβ (+) controls ( $p < 0.05$ )
<sup>e</sup> Significant difference between Aβ (-) patients and Aβ (+) controls ( $p < 0.05$ )
<sup>f</sup> Significant difference between Aβ (-) patients and Aβ (-) controls ( $p < 0.05$ )
Key: HAB, high affinity binder; MAB, mixed affinity binder; MMSE, Mini-Mental Status
Examination; SRT-DR, Selective Reminding Test-Delayed Recall; TSPO, 18 kDa translocator protein.

Among all included participants who had UPSIT, amyloid-positive patients and amyloid-negative controls were younger than amyloid-positive controls and amyloid-negative patients (*p <* 0.01). We found no difference in years of education among participant groups. Amyloid-positive patients had lower MMSE scores than amyloid-negative controls, amyloid negative patients and amyloid positive controls (*p’s* < 0.01). Both amyloid-positive patients and amyloid-negative patients had smaller hippocampal volume, lower SRT-DR scores, and lower MMSE scores than the control groups (*p <* 0.01), suggesting that the impaired participants had hippocampal atrophy even when amyloid pathology was absent. There were more men in the amyloid-positive and amyloid-negative patient groups and more women among the amyloid-negative controls (χ² (3, *N* = 54) = 10.9, *p =* 0.012), so statistical analysis accounted for sex as a co-variate.

### UPSIT performance across study groups

We tested whether amyloid status and cognitive status were independently associated with UPSIT performance. We found that amyloid positivity (*F*_1, 48_= 9.15, *p* = 0.004) and cognitive impairment (*F*_1, 48_ = 8.66, p = 0.005) were each negatively associated with UPSIT score. These associations remained after controlling for age, sex and *TSPO* genotype. However, we found no interaction between amyloid status and cognitive status (*F*_1, 47_ = 0.776, *p* = 0.383). The T of amyloid status and cognitive status were 0.103 and 0.163, respectively.

Amyloid-positive patients had lower UPSIT scores than amyloid-negative controls (*p* < 0.01, Fig 1). UPSIT performance of amyloid-negative patients did not differ significantly from amyloid-negative controls (p = 0.13) or amyloid-positive patients (*p* = 0.97). The 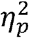 of participant groups was 0.330.

**Figure 1.**
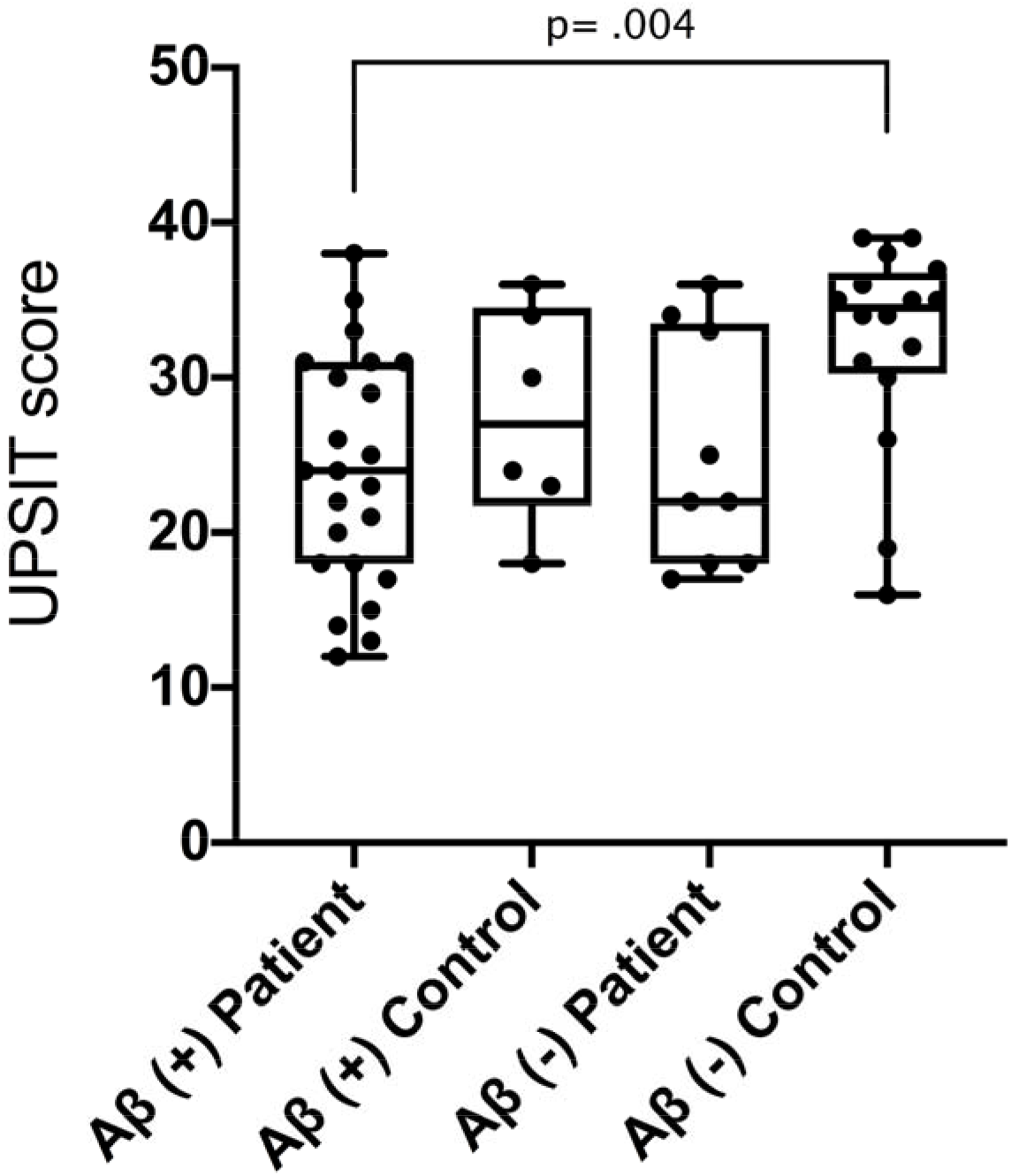
UPSIT performance across study groups. UPSIT scores across all four study groups. UPSIT scores were lower in amyloid-positive patients than amyloid-negative controls.

### UPSIT performance and ^18^F-MK-6240 binding

For participants who underwent ^18^F-MK6240 PET imaging (n= 41), we performed a partial correlation analysis between ^18^F-MK-6240 binding and UPSIT performance. Using the partial volume corrected SUVR data, we found that ^18^F-MK-6240 binding was negatively associated with UPSIT performance in all ROIs except lingual gyrus when all participants were combined (*r*’s > −0.35, *p’s* < 0.05) (Fig 2, Table 2). Correlations between UPSIT performance and ^18^F-MK6240 binding in all ROIs except for the lingual gyrus remained significant after multiple comparison correction (Table 2). When we stratified participants based on amyloid status, this significant negative partial correlation remained for amyloid-positive participants in medial temporal cortex (*r_p_* = −0.51, *p* = 0.02) and hippocampus (*r_p_* = −.53, *p* = 0.02). ^18^F-MK-6240 binding did not correlate with UPSIT performance in any regions when only amyloid-negative participants were included, not even at trend level (e.g., medial temporal cortex (*r_p_* = −0.27, p = 0.31), hippocampus (*r_p_* = −.17, *p* = 0.54)). Results from correlation analysis using PET data uncorrected for partial volume effects showed similar results (Supplementary Table 1).

**Table 2.** Correlation analysis between UPSIT and partial volume corrected ^18^F-MK6240 binding.

| Region of Interest | All participants (n = 41) | | | Aβ (+) participants<br>(n = 22) | | | Aβ (-) participants<br>(n = 19) | | |
| --- | --- | --- | --- | --- | --- | --- | --- | --- | --- |
|  | r | 95%<br>CI | P-value | r | 95%<br>CI | P-value | r | 95%<br>CI | P-value |
| Pre-frontal Cortex | -0.44* | -0.66–<br>-0.16 | 0.005 | -0.37 | -0.69–<br>0.06 | 0.12 | -0.32 | -0.68–<br>0.16 | 0.23 |
| Middle and Inferior<br>Temporal Gyri | -0.48* | -0.69–<br>-0.20 | 0.002 | -0.38 | -0.69–<br>0.05 | 0.11 | -0.24 | -0.63–<br>0.24 | 0.37 |
| Superior Temporal<br>Cortex | -0.43* | -0.65–<br>-0.14 | 0.006 | -0.32 | -0.65–<br>0.12 | 0.19 | -0.18 | -0.59–<br>0.30 | 0.51 |
| Medial Temporal<br>Cortex Composite | -0.59* | -0.76–<br>-0.35 | 0.00 | -0.52 | -0.77–<br>-0.12 | 0.02 | -0.25 | -0.63–<br>0.23 | 0.36 |
| Posterior Cingulate<br>Cortex | -0.35* | -0.60–<br>-0.05 | 0.03 | -0.14 | -0.53–<br>0.30 | 0.56 | -0.14 | -0.56–<br>0.34 | 0.60 |
| Superior Parietal<br>Cortex | -0.36* | -0.60–<br>-0.06 | 0.02 | -0.18 | -0.56–<br>0.26 | 0.46 | -0.04 | -0.48–<br>0.42 | 0.89 |
| Inferior Parietal<br>Cortex | -0.39* | -0.62–<br>-0.09 | 0.02 | -0.20 | -0.57–<br>0.25 | 0.42 | -0.11 | -0.54–<br>0.37 | 0.69 |
| Striatum | -0.35* | -0.60–<br>-0.05 | 0.03 | -0.08 | -0.48–<br>0.36 | 0.76 | -0.29 | -0.66–<br>0.18 | 0.27 |
| Hippocampus | -0.60* | -0.76– | 0.00 | -0.53 | -0.78– | 0.02 | -0.27 | -0.65– | 0.31 |
|  |  | -0.36 |  |  | -0.14 |  |  | 0.21 |  |
| Lingual Gyrus | -0.21 | -0.49–<br>0.10 | 0.20 | 0.02 | -0.40–<br>0.44 | 0.92 | -0.02 | -0.47–<br>0.44 | 0.94 |

\* Survived multiple comparison correction.

**Figure 2.**
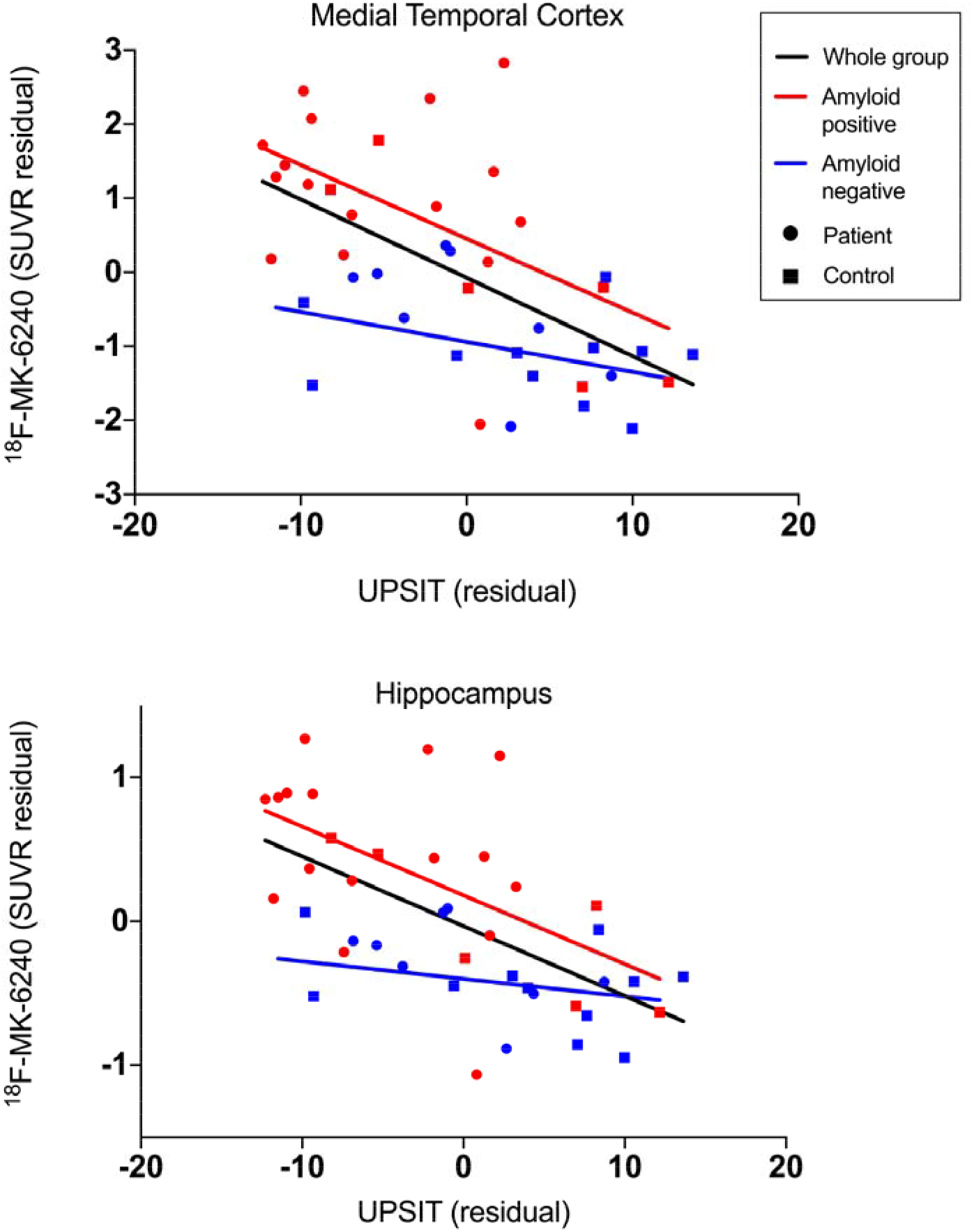
Relationship between UPSIT score and ^18^F-MK620 PET. Lower UPSIT scores were associated with greater ^18^F-MK-6240 binding when all participants included (medial temporal cortex (r = −0.59, *p* < 0.01) and hippocampus (r = −0.60, *p* < 0.01). Correlations remained when only amyloid-positive participants were included (medial temporal cortex: r = −0.52, *p* = 0.02; hippocampus: r = −0.53, p = 0.02) but not when only amyloid-negative participants were included. Data corrected for age and sex.

### UPSIT performance and ^11^C-PBR28 binding

For participants who underwent ^11^C-PBR28 PET imaging (n= 53), we performed a partial correlation analysis between ^11^C-PBR28 binding and UPSIT performance. Using the partial volume corrected SUVR data, we found that ^11^C-PBR28 binding was negatively associated with UPSIT performance in the middle and inferior temporal gyri, medial temporal cortex, posterior cingulate cortex, inferior parietal cortex and hippocampus when all participants were combined (*r_p_’s* > −0.29, *p’s* < 0.05, Fig 3, Table 3). Correlations between UPSIT performance and ^11^C-PBR28 binding in the medial temporal cortex, posterior cingulate cortex, hippocampus and middle and inferior temporal gyri remained significant after multiple comparison correction (Table 3). When we stratified participants based on amyloid status, this negative partial correlation remained for amyloid-positive participants in the medial temporal cortex (*r_p_* = −0.74, *p < 0.001*) and the middle and inferior temporal gyri (*r_p_* = −0.48, *p* = 0.02). When only amyloid-negative participants were included, ^11^C-PBR28 binding did not correlate with UPSIT performance in any region except at trend level for the hippocampus (*r_p_* = −0.38, *p* = 0.08). Results from partial correlation analysis using imaging data uncorrected for partial volume effects showed similar results (Supplementary Table 2).

**Table 3.** Correlation analysis between UPSIT and partial volume corrected ^11^C-PBR28 binding.

|  | **All participants (n = 53)** |  |  | **Aβ (+) participants**<br>**(n = 28)** |  |  | **Aβ (-) participants**<br>**(n = 25)** |  |  |
| --- | --- | --- | --- | --- | --- | --- | --- | --- | --- |
| **Region of Interest** | **r** | **95%**<br>**CI** | **P-**<br>**value** | **r** | **95%**<br>**CI** | **P-value** | **r** | **95%**<br>**CI** | **P-**<br>**value** |
| Pre-frontal Cortex | -0.22 | -0.46–<br>0.05 | 0.12 | -0.19 | -0.53–<br>0.20 | 0.36 | 0.12 | -0.28–<br>0.50 | 0.58 |
| Middle and<br>Inferior Temporal<br>Gyri | -0.47* | -0.66– -<br>0.23 | 0.00 | -0.47 | -0.72–<br>-0.12 | 0.02 | -0.09 | -0.47–<br>0.031 | 0.68 |
| Superior Temporal<br>Cortex | -0.20 | -0.45–<br>0.07 | 0.16 | -0.15 | -0.50–<br>0.24 | 0.48 | 0.16 | -0.25–<br>0.52 | 0.49 |
| Medial Temporal<br>Cortex Composite | -0.58* | -0.74– -<br>0.37 | 0.00 | -0.74 | -0.87–<br>-0.50 | 0.00 | -0.06 | -0.45–<br>0.34 | 0.77 |
| Posterior Cingulate<br>Cortex | -0.34* | -0.56– -<br>0.08 | 0.01 | -0.15 | -0.49–<br>0.24 | 0.48 | -0.18 | -0.54–<br>0.23 | 0.43 |
| Superior Parietal<br>Cortex | -0.25 | -0.49–<br>0.02 | 0.08 | 0.00 | -0.34–<br>0.38 | 0.98 | 0.10 | -0.31–<br>0.47 | 0.67 |
| Inferior Parietal<br>Cortex | -0.29 | -0.52– -<br>0.02 | 0.04 | -0.15 | -0.50–<br>0.23 | 0.46 | 0.10 | -0.31–<br>0.47 | 0.67 |
| Striatum | -0.09 | -0.36–<br>0.18 | 0.51 | -0.07 | -0.44–<br>0.31 | 0.72 | -0.15 | -0.51–<br>0.26 | 0.50 |
| Hippocampus | -0.35* | -0.57– - | 0.01 | -0.31 | -0.61– | 0.15 | -0.38 | -0.67– | 0.08 |
|  |  | 0.08 |  |  | 0.08 |  |  | 0.02 |  |
| Lingual Gyrus | -0.20 | -0.45–<br>0.07 | 0.16 | -0.02 | -0.39–<br>0.36 | 0.94 | 0.06 | -0.34–<br>0.44 | 0.79 |

\*Survived multiple comparison correction

**Figure 3.**
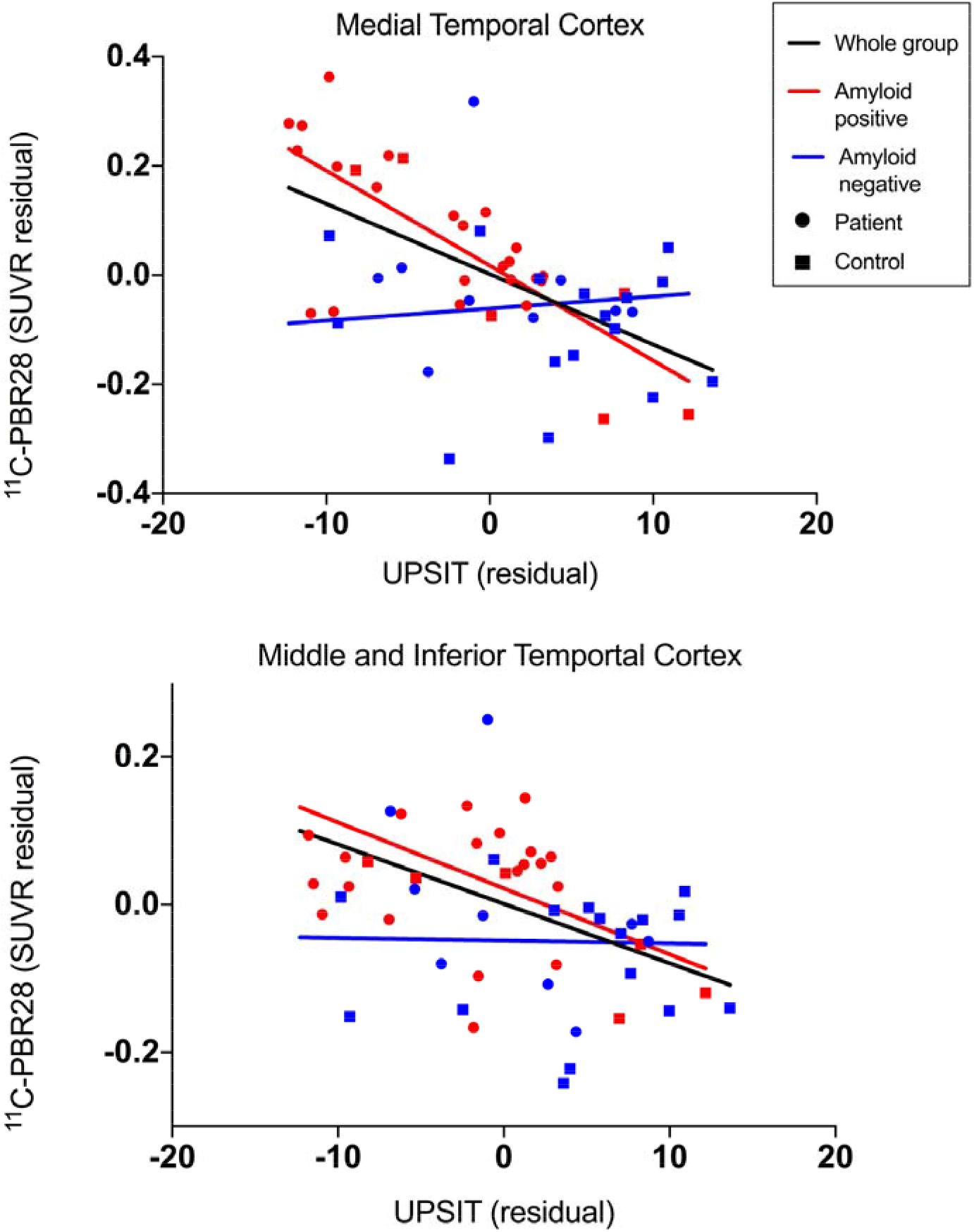
Relationship between UPSIT score and ^11^C-PBR28 PET. Lower UPSIT scores were associated with greater ^11^C-PBR28 binding when all participants were included in medial temporal cortex (r = −0.58, *p* < 0.01) and combined middle and inferior temporal gyri (r = −0.47, *p* < 0.01). Correlations remained when only amyloid-positive participants were included (medial temporal cortex: r = −0.74, *p* < 0.01; combined middle and inferior temporal gyri: r = −0.47, p = .02) but not when only amyloid-negative participants were included. Data corrected for age, sex, and *TSPO* genotype.

### UPSIT performance and CSF biomarkers

For participants who underwent lumbar puncture (n=23), we performed a partial correlation analysis between CSF biomarkers burden and UPSIT performance. We found that UPSIT performance was negatively associated with CSF concentrations of t-tau (*r_p_* = −0.52, p = 0.02) and p-tau (*r_p_* = −0.53, p = 0.012) when all participants were combined (Fig 4, Table 4). We did not observe a significant negative association between UPSIT performance and CSF concentrations of Aβ_42_ (*r_p_* = −0.12, p = 0.60). Correlations between UPSIT performance and CSF measures of t-tau and p-tau remained significant after multiple comparison correction (Table 4). Due to the smaller sample size of participants who underwent lumbar puncture, we did not stratify participants based on amyloid status for subgroup evaluation.

**Table 4.** Correlation analysis between UPSIT and CSF measures

| **CSF Biomarker** | **r (whole Group)** | **95% CI** | **P-value** |
| --- | --- | --- | --- |
| T-tau | -0.52 | -0.77– -0.14 | 0.02 |
| P-tau | -0.53 | -0.78– -0.16 | 0.02 |
| Aβ<sub>42</sub> | 0.14 | -0.30– 0.51 | 0.60 |

\* Survived multiple comparison correction

**Figure 4.**
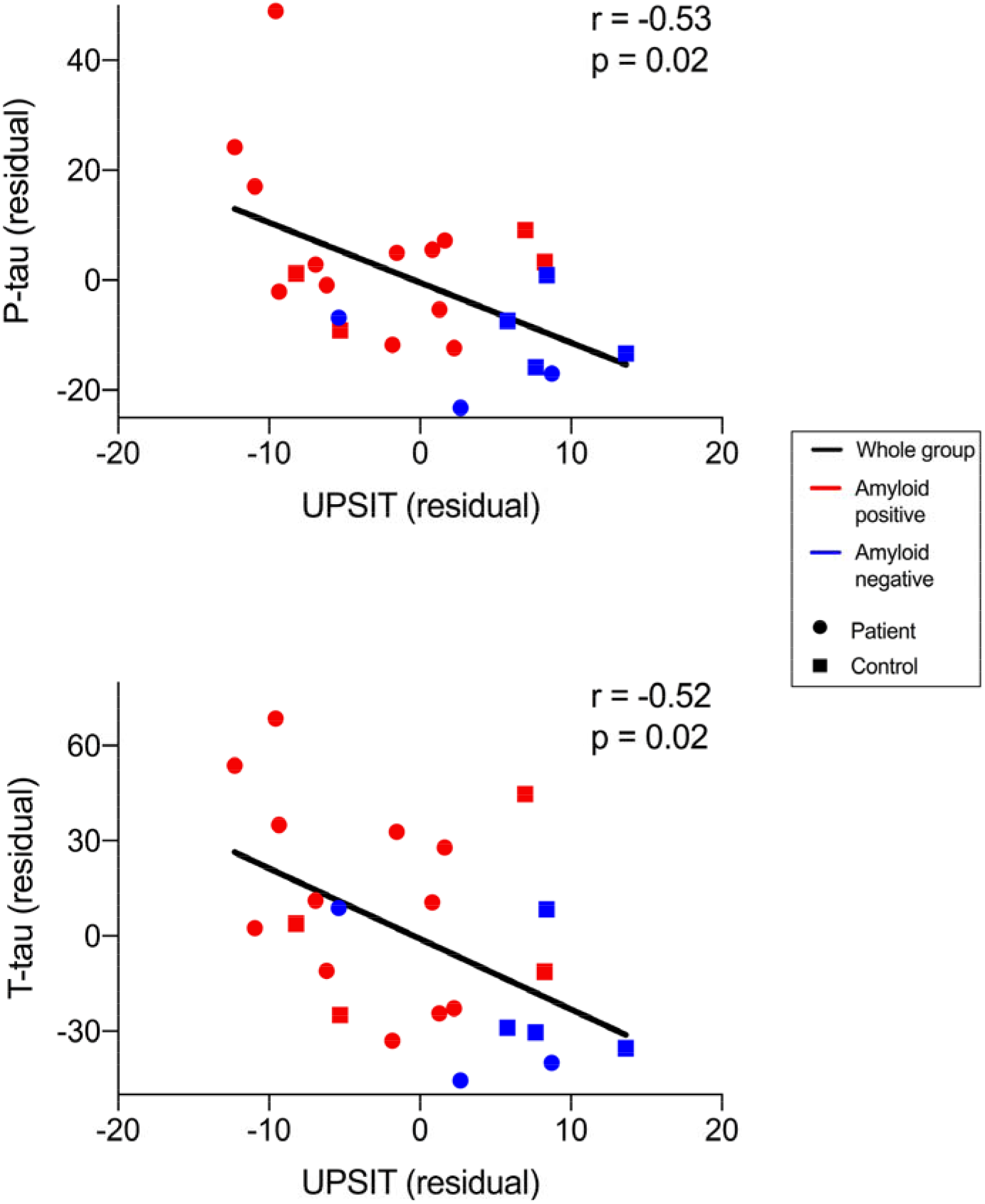
Relationship between UPSIT score and CSF concentrations of total tau and phosphorylated tau. Lower UPSIT scores were associated with greater CSF concentrations of phosphorylated tau (p-tau, r = −0.53, *p* = .02) and total tau (t-tau, r = −0.52, *p* = .02), after controlling for age and sex.

### UPSIT performance, hippocampal volume, and cognition

Performance on the UPSIT positively correlated with hippocampal volume, such that lower UPSIT scores were associated with smaller hippocampal volumes, when all participants were included (*r_p_* = 0.53, *p* < 0.001) and when only amyloid-positive participants were included (*r_p_* = 0.69, *p* < 0.001, Fig 5A). UPSIT performance positively correlated with MMSE scores (*r_p_* = 0.42, *p* < 0.001) and z-scores for performance on the SRT-DR (*r_p_* = 0.65, *p* < 0.001), such that lower UPSIT scores were associated with worse cognitive performance, when all participants were included (Fig 5B). The partial correlation between UPSIT and SRT-DR performance remained significant when only amyloid-positive participants were included (*r_p_* = 0.68, *p* < 0.001, Fig 5C).

**Figure 5.**
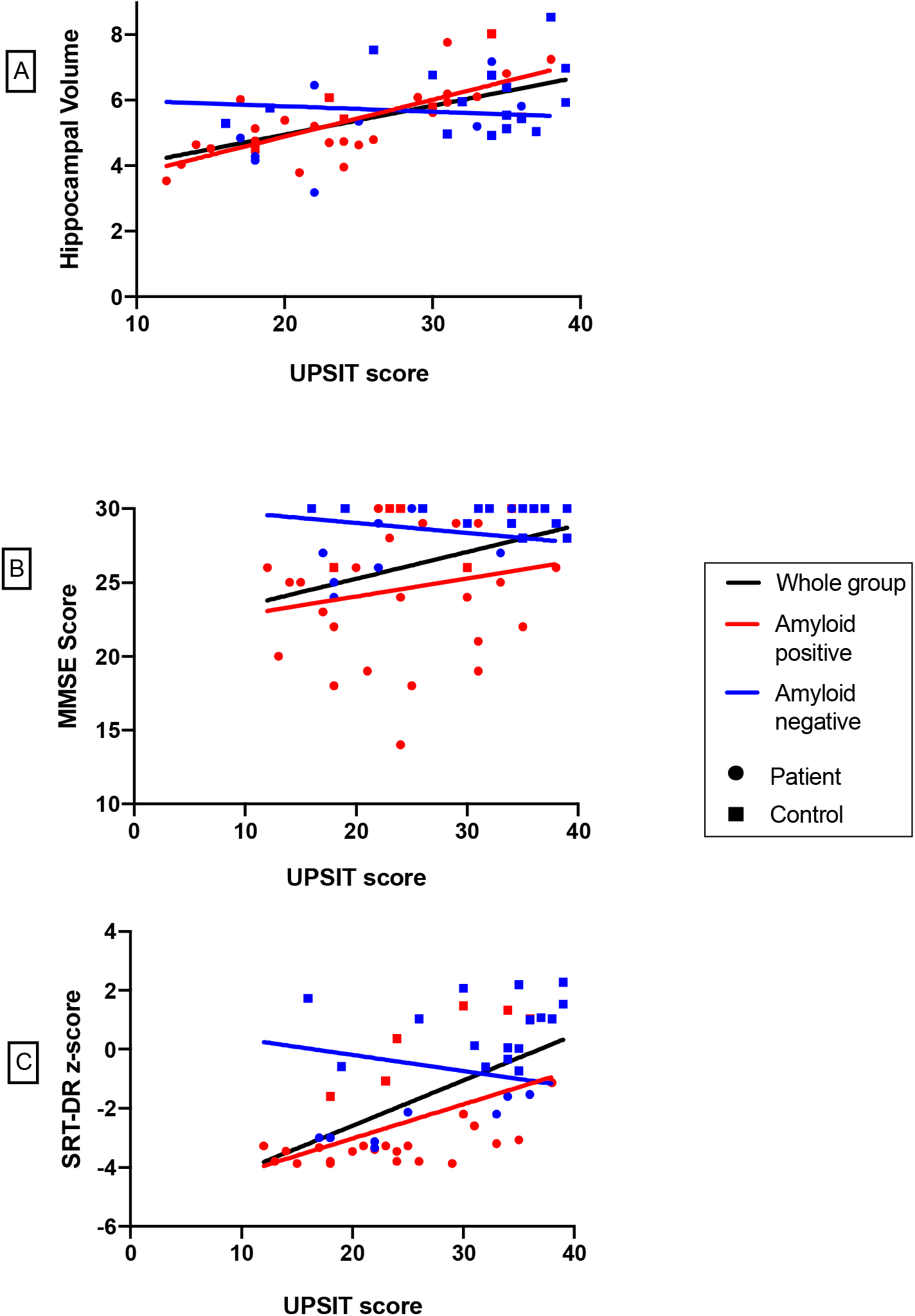
Relationship among UPSIT score and hippocampal volume, Mini Mental State Exam (MMSE) score and Selective Reminding Test – Delayed Recall (SRT-DR) score. Positive correlations were observed between UPSIT performance and (A) hippocampal volume (*r* = 0.53, *p*< 0.001), (B) MMSE (*r* = 0.42, *p*< 0.001) and (C) SRT-DR performance (*r* = 0.65, *p*< 0.001) when all participants were included. Positive correlations between UPSIT performance and hippocampal volume (*r*= 0.69, *p*< 0.001) and UPSIT and SRT-DR performance (*r* = 0.68, *p*< 0.001) remained when only amyloid-positive participants were included.

## DISCUSSION

We demonstrated that olfactory identification is negatively associated with progression along the Alzheimer’s disease clinical continuum, such that amyloid-positive patients had lower UPSIT scores than amyloid-negative controls, and that UPSIT score positively correlated with cognitive performance and hippocampal volume. We also found that UPSIT score negatively correlated with PET and CSF measures of tau pathology and neuroinflammation. Taken together, these results suggest that odor identification worsens with AD progression in a manner that may be related to both tau and neuroinflammatory burden.

When we considered the amyloid-positive group separately, we found inverse relationships between olfactory identification ability and both tau pathology and neuroinflammation in medial temporal regions (hippocampus and the combined amygdala/parahippocampal gyrus). This topographical specificity is notable because these regions, which are affected early in AD, receive afferent input from primary neurons originating in the olfactory bulb [5]. Our results suggest that decreased ability to identify odors may reflect the burden of tau-mediated neurodegeneration in early Braak regions, and UPSIT may have utility in the early detection of AD pathophysiology.

Our results build on early findings demonstrating relationships between UPSIT performance and AD. In one sample of cognitively normal adults, poorer performance on the 12-item Brief Smell Identification (B-SIT) was associated with a 50% increased risk of developing MCI over the following five years and exhibited predictive value for developing dementia [12]. Lower UPSIT scores are associated with smaller hippocampal and entorhinal volumes in cognitively normal elders, particularly in those with high amyloid burden on PET [9, 10]. Another PET study found that lower odor identification scores in a shorter version of the UPSIT were modestly associated with greater neocortical amyloid binding in cognitively normal, MCI and AD patients when combined, but not when MCI participants were considered independently, suggesting that olfactory impairment is not directly related to amyloid burden alone [29]. Additionally, UPSIT performance prior to death predicted neurofibrillary tangle burden in the CA1/subiculum of the hippocampus in AD patients [30]. Our finding of a negative relationship between UPSIT score and CSF concentrations of tau, but not Aβ_42_ is in agreement with prior studies showing that low performance on the B-SIT and UPSIT have been associated with increased CSF tau, increased CSF t-tau: Aβ_42_ ratios and increased p-tau (181): Aβ_42_ ratios but not with measurement of CSF Aβ_42_.[14, 31] These results suggest that UPSIT may provide more insight into burden of tau pathology in early AD than amyloid pathology.

To our knowledge, the only prior study comparing odor identification and tau pathology *in vivo* using PET imaging used Flortaucipir (^18^F-AV-1451) and likewise found an inverse relationship between UPSIT score and tau binding in temporal and parietal cortices in cognitively normal adults and patients with subjective cognitive decline. In our study, we extended these results to include clinically affected AD patients and patients who are amyloid-negative but exhibit evidence of hippocampal neurodegeneration and AD patterns of cognitive impairment. In addition, we used ^18^F-MK-6240, an improved tau radioligand with less off-target binding in basal ganglia and choroid plexus than ^18^F-AV-1451, and confirmed our imaging findings by demonstrating correlations between UPSIT score and CSF concentrations of t-tau and p-tau [25, 32].

We also showed that there is a strong relationship between UPSIT performance and PET measures of neuroinflammation. While the staging of neuroinflammation is poorly understood, one meta-analysis including results from a range of TSPO radioligands, including ^11^C-PBR28, reported that the difference in microglial activation measured on PET imaging between AD patients and healthy controls existed in several cortical areas, but was greatest in the middle and inferior temporal gyri and the parahippocampal gyrus [33]. The same relationships were observed in MCI, albeit more modestly. Interestingly, these regions were among those that exhibited the strongest inverse relationships with UPSIT performance in our present study, suggesting that these regions experience early neuroinflammation in AD. Further, the inverse relationships between UPSIT performance and PET measures of neuroinflammation were observed in nearly identical brain regions as PET measures of tau pathology. These results support the possibility of a topographical overlap between neuroinflammation and tau deposition in early neurodegeneration, and align with results of prior PET studies showing colocalization of TSPO and tau binding [34, 35]. Therefore, neuroinflammation and tau pathology appear to be closely related across the AD continuum and may both be contributing factors in the development of olfactory impairment.

Our current study selected a subset participants from a pre-established research cohort. In a prior study of the larger cohort, both ^18^F-MK-6240 and ^11^C-PBR28 binding were greater in amyloid-positive than in amyloid-negative participants, specifically in neocortical regions for ^11^C-PBR28 and in the medial temporal lobe for ^18^F-MK-6240 [35]. In our sub-sample, we observed similar regional patterns of increased ^11^C-PBR28 and ^18^F-MK-6240 binding in association with lower UPSIT scores, suggesting that odor identification impairment may be mechanistically linked to inflammation and tau pathology, and not just a nonspecific measure of neurodegeneration.

We also found that amyloid status and cognitive status are independently associated with UPSIT performance. That amyloid-positivity is associated with lower UPSIT score is consistent with prior studies showing that lower performance on odor identification predicts decline in cognitively normal elderly and UPSIT scores correlate with amyloid deposition on PET [36, 37]. Therefore, UPSIT may be useful as a selection tool to identify cognitively normal elders more likely to be amyloid-positive for preventative clinical trials. That impaired cognition is associated with lower UPSIT score independent of amyloid status is not surprising, given that impaired odor identification has also been reported in amyloid negative dementias, or non-AD dementias such as dementia with Lewy bodies, Huntington’s disease, and frontotemporal dementia [38]. Notably, amyloid-negative patients did not have lower UPSIT scores than amyloid-negative controls, although this may relate to our modest sample size. That amyloid-positive patients had the lowest UPSIT scores in our cohort may reflect greater overall pathology in this group than in the amyloid-negative participants. We don’t have histopathological confirmation in the amyloid-negative patients; however, given the overall small hippocampal volumes and AD-like patterns of impairment in this group, they may represent hippocampal sclerosis/TDP-43 pathology, argyrophilic grain disease, or other AD mimics that may have more indolent clinical trajectories than patients with biomarker evidence of AD pathophysiology [39]. Importantly, none of the amyloid-negative patients had clinical or radiographic (on MRI) signs or symptoms indicative of non-AD dementias such as frontotemporal dementia, dementia with Lewy bodies, vascular dementia, progressive supranuclear palsy, or corticobasal syndrome).

Our conclusions are limited by our sample size. We did not observe any significant relationships within the amyloid-negative subgroups alone and many of the overall relationships observed were driven by amyloid-positive patients. We cannot say, however, that olfactory identification is not related to tau or neuroinflammation in amyloid-negative participants, only that we failed to find such a relationship and that presumably tau, neuroinflammation, and impaired odor identification are mediated at least in part by amyloid. Sample size was particularly limiting for our CSF analysis, as only 23 participants in our cohort elected to have lumbar puncture performed, and therefore we did not evaluate amyloid-positive and amyloid-negative groups separately. Further, our sample size did not permit stratification of these relationships by sex given that there were more male participants in both the amyloid-positive and amyloid-negative patient groups and more female participants overall in the control group. However, sex was considered as a biological covariate in statistical analysis. We did not evaluate relationships between olfactory impairment and performance on neuropsychological testing batteries beyond the MMSE and SRT-DR. However, the relationship between UPSIT and cognitive performance was more comprehensively investigated in a larger community cohort of over 1000 participants, with results demonstrating that UPSIT significantly correlated with neuropsychological measures of memory, fluency and executive functioning [12]. While these results do not imply causation due to the limitations of a cross-sectional observational study, our results indicate that there appear to be significant associations between olfactory impairment, tau pathology and neuroinflammation that could be further investigated with a larger sample size. ^18^F-MK-6240 is still an early tau radioligand with an off-target binding profile that is not yet fully understood. Early studies, however, suggest that the radioligand has adequate sensitivity for detecting tau pathology [32, 40].

In conclusion, while reduced olfactory identification ability has previously been linked to cognitive decline and amyloid deposition, we have demonstrated that UPSIT performance is also related to other contributors of AD pathophysiology. Therefore, the UPSIT appears to serve broader utility beyond being a marker of disease severity, but rather an inexpensive, non-invasive screening tool that may provide insight into the burden of tau pathology and neuroinflammation. Based on our results and the literature, the UPSIT could also be considered for use as an initial screening tool to identify participants in at-risk populations who may be more likely to test positive of PET for amyloid, tau or other *in vivo* measures of AD pathology, saving time and cost in clinical trials involving preventative treatments.

## Data Availability

Anonymized data will be made available upon reasonable request to qualified investigators. William C. Kreisl takes full responsibility for the data, the analyses and interpretation, and the conduct of the research. He has full access to all of the data and has the right to publish any and all data separate and apart from any sponsor.

## ACKNOWLEDGEMENTS

^18^F-Florbetaben was supplied by Life Molecular Imaging. ^18^F-MK-6240 was supplied by Cerveau Technologies. *TSPO* genotyping was performed by Regina Santella, PhD and the Columbia University Biomarkers Shared Resource. We wish to acknowledge the contributions of the staff and faculty of the Irving Institute for Clinical Research, the MRI Center and the David A. Gardner PET Imaging Center at the Columbia University Irving Medical Center.

